# Association of genetic variants from the Wnt signaling pathway with cardiovascular disease in the Saudi Arabian population

**DOI:** 10.64898/2026.08.21.26361030

**Authors:** CR Kaundinya, Narasimha Reddy Parine, Maha Arafah, Jilani Purusottapatnam Shaik, Akbar Ali Khan Pathan

## Abstract

The canonical Wnt/β-catenin signaling pathway plays a key role in cardiovascular development, preservation, and pathology. Variations in critical Wnt pathway genes may influence an individual’s susceptibility to cardiovascular disease (CVD), although data from specific populations are scarce. In this case–control study, we analyzed 15 single-nucleotide polymorphisms (SNPs) within eight Wnt pathway genes (APC, AXIN2, LRP6, CTNNB1, TCF7L2, DKK3, DKK4, and SFRP3) among 151 CVD patients and 129 healthy controls. We examined the genotypic and allelic distributions for correlations with CVD risk utilizing odds ratios, confidence intervals, and chi-square tests, while controlling for age and gender. We discovered that the APC variants rs459552 and rs454886 conferred protective effects, with age-and gender-dependent variation. AXIN2 SNP rs11079571 made men more likely to get CVD, and rs3923086 made people over 58 more susceptible. The DKK4 variant rs3763511 was associated with an elevated risk of cardiovascular disease, particularly among males and older individuals (age M/F). In SFRP3, rs7775 was associated with an elevated risk in older individuals (age M/F), whereas rs288326 showed a protective effect. For LRP6, rs2284396 increased the risk of CVD in females, while rs2075241 conferred protection in males. We did not identify significant associations for the CTNNB1, TCF7L2, or DKK3 variants. The present data indicate that specific Wnt pathway variants are associated with cardiovascular disease risk, contingent on age and gender. To verify these outcomes and determine whether these variants can serve as genetic markers of cardiovascular disease risk, larger, more diverse studies with whole genome sequencing approach is necessary.

## Introduction

The Wnt signaling pathway is a preserved and essential mechanism for intercellular communication. It regulates diverse biological processes, including cell fate specification, tissue organization, proliferation, migration, and environmental response (Clevers 2013; Logan and Nusse 2004). Dysregulation of Wnt signaling, active from embryogenesis to adulthood, is involved in developmental, degenerative, and metabolic disorders (Nusse and Clevers 2017).

Wnt ligands are secreted glycoproteins that are modified in the endoplasmic reticulum. These proteins are transported to the cell surface via Wntless (Nusse and Clevers 2017; Xue et al. 2025), where Wnt signaling initiates. Wnt proteins bind mainly to Frizzled receptors, which are seven-transmembrane proteins (Nusse and Clevers 2017; Xue et al. 2025), and affect nearby cells. In the presence of LRP5 or LRP6 co-receptors, the main β-catenin–dependent pathway is activated (Nusse and Clevers 2017; Xue et al. 2025). Other receptors, such as ROR1, ROR2, or RYK, can activate distinct Wnt pathways independent of β-catenin (Nusse and Clevers 2017; Xue et al. 2025). This discussion does not focus on these alternative pathways.

Cells are in a Wnt-off state when Wnt ligands are not present. This means that they tightly control how much β-catenin is inside the cell. Even though β-catenin is always being made, a group of proteins, such as APC, AXIN, GSK-3β, and CK1, quickly breaks it down (Nusse and Clevers 2017; Xue et al. 2025). AXIN helps organize the process so that CK1 and GSK-3β can add phosphate groups to β-catenin (Nusse and Clevers 2017; Xue et al. 2025). The cell’s proteasome system degrades β-catenin after it is phosphorylated (Nusse and Clevers 2017; Xue et al. 2025). In this state, TCF/LEF transcription factors remain bound to DNA but also interact with repressors, so Wnt target genes are not turned on (Nusse and Clevers 2017; Xue et al. 2025).

When Wnt ligands bind to Frizzled receptors and LRP5/6 co-receptors, cell signaling changes immediately (Liu et al. 2022; Nusse and Clevers 2017; Xue et al. 2025). This binding enables the protein disheveled to function as a key signaling messenger (Liu et al. 2022; Nusse and Clevers 2017; Xue et al. 2025). Dishevelled protein then adds phosphate groups to LRP5/6 and moves AXIN to the cell membrane, which breaks up the β-catenin destruction complex (Liu et al. 2022; Nusse and Clevers 2017; Xue et al. 2025). This causes β-catenin to accumulate in the cell and enter the nucleus. There, β-catenin removes repressors from TCF/LEF complexes and recruits coactivators such as CBP and p300. This makes TCF/LEF activators and turns on Wnt target genes (Liu et al. 2022; Nusse and Clevers 2017; Xue et al. 2025).

Wnt ligands can initiate non-canonical Wnt pathways independent of β-catenin. Small GTPases such as RhoA and Rac are used by the planar cell polarity pathway to control cell orientation (Butler and Wallingford 2017). The Wnt/Ca²⁺ pathway increases intracellular calcium levels and activates enzymes such as protein kinase C, CaMKII, and calcineurin. These enzymes then alter how cells stick together, move, and alter gene expression in response to the situation.

Wnt signaling is important for more than just embryonic development; it also helps renew tissues in organs such as the skin, intestine, and bone (Gough 2012). Moreover, it helps control stem cells after they have grown and keeps a balance between cell growth and specialization (Gough 2012; Liu et al. 2022; Nusse and Clevers 2017; Xue et al. 2025). If this balance is not kept, it can lead to illness. For instance, mutations in APC or β-catenin that keep Wnt signaling going all the time are linked to a number of diseases (Xue et al. 2025). Furthermore, abnormal Wnt signaling is now known to be involved in chronic and degenerative diseases, in addition to cancer, including CVD (Tian et al. 2010).

Coronary artery disease, stroke, peripheral artery disease, heart failure, arrhythmias, valve problems, and birth defects are all types of CVD. These diseases are a major cause of illness and death worldwide (Roth et al. 2020). The World Health Organization estimates that CVD causes about 17.9 million deaths every year. This is nearly a third of all deaths globally (World Health Organization, n.d.).

CVD develops and worsens due to environmental, lifestyle, and genetic factors (Yusuf et al. 2020). Common risk factors include high blood pressure, high cholesterol, smoking, being overweight, diabetes, inactivity, genetics, and older age (Yusuf et al. 2020). Genetic variations can affect disease onset, progression, and treatment response (Inouye et al. 2018; Kathiresan and Srivastava 2012). CVD can result from atherosclerosis, abnormalities of the blood vessel lining, chronic inflammation, oxidative stress, abnormal fat metabolism, vascular smooth muscle cell dysfunction, or harmful structural changes in the heart (Bentzon et al. 2014; Gimbrone and García-Cardeña 2016; Owens et al. 2004).

Several signaling pathways, including Wnt, PI3K/Akt, MAPK/ERK, JAK/STAT, NF-κB, TGF-β, and calcium signaling, help the heart and blood vessels function properly (Hayden and Ghosh 2011; Johnson and Lapadat 2002; Manning and Toker 2017; Nusse and Clevers 2017; O’Shea and Plenge 2012). Among these, Wnt signaling is crucial for heart and blood vessel development in embryos (Cohen et al. 2007). In adults, Wnt signaling is predominantly inactive under normal circumstances but reactivates in response to stress or injury (Aisagbonhi et al. 2011; Bergmann 2010; Shimizu and Minamino 2016).

The principal β-catenin–dependent Wnt pathway is key in the pathogenesis of CVD. Reactivating β-catenin signaling can cause heart tissue scarring, thickening, problems with the vessel lining, changes in blood vessels, calcium buildup, and poor heart function (Bergmann 2010; Bundy et al. 2021; Gimbrone and García-Cardeña 2016; Shimizu and Minamino 2016; Stylianidis et al. 2016; Zhou et al. 2008). Wnt signaling turns cardiac fibroblasts into myofibroblasts after heart injury. This process creates excess matrix, making the heart stiffer (Zhou et al. 2008). Continued β-catenin activation in cardiac myocytes can cause harmful hypertrophy and increase the risk of arrhythmias and cardiac failure (Bergmann 2010; Shimizu and Minamino 2016; Stylianidis et al. 2016). In blood vessels, abnormal Wnt signaling disrupts the lining, increases smooth muscle proliferation, alters cell types, promotes plaque buildup, and leads to arterial calcification, especially in those with diabetes or renal disease (Bundy et al. 2021; Mill and George 2012; Shroff et al. 2013).

Even though treatments like β-blockers and calcium channel blockers have gotten better, it’s still hard to manage CVD. This is partly because genetic differences between people affect their risk of CVD and their response to treatment (Khot et al. 2003; Torkamani et al. 2018). CVD can also go undetected for years before suddenly leading to a heart attack or stroke. This shows how important it is to improve early screening and genetic risk assessment.

Canonical Wnt signaling plays a crucial role in heart development, maintenance, and disease, making it a strong candidate for genetic research and biomarker discovery. Genetic changes in APC, AXIN2, β-catenin, LRP6, and TCF4 can cause Wnt signaling to persistently or abnormally remain active, leading to sustained activation of β–catenin–dependent transcription even in the absence of Wnt ligands ((Dai et al. 2019; Mani et al. 2007). Such abnormal activation is linked to cancer and various diseases, including cardiovascular disease (Xue et al. 2025).

This study investigated genetic variants in essential genes of the canonical Wnt signaling pathway and evaluated their correlation with cardiovascular disease in the Saudi Arabian population. Notably, genome-wide association studies (GWAS) have demonstrated that single-nucleotide polymorphisms (SNPs) significantly influence the genetics of cardiovascular diseases. Based on these findings, we selected SNPs from the SNP500Cancer project that map to genes in the Wnt pathway.

Fifteen SNPs in eight genes associated with the Wnt signaling pathway, potentially influencing cardiovascular disease susceptibility, were examined. We examined changes in APC, AXIN2, β-catenin (CTNNB1), LRP6, and TCF4, as well as canonical Wnt modulators such as DKK3, DKK4, and the secreted Wnt inhibitor SFRP3. These variants were examined in 151 individuals diagnosed with cardiovascular disease. We examined 15 SNPs within eight Wnt pathway genes that may influence cardiovascular disease risk. These encompassed variants in APC, AXIN2, β-catenin (CTNNB1), LRP6, TCF4, and Wnt modulators such as DKK3, DKK4, and SFRP3. The research involved 151 individuals with CVD and 129 healthy controls to investigate the potential contribution of these variants to the disease and their efficacy as genetic markers for CVD risk. By examining the distribution and functional relevance of these variants, this study aims to identify potential biomarkers associated with CVD and to enhance our understanding of the genetic architecture underlying cardiovascular disease susceptibility. Example, APC rs459552 could help with early screening, and AXIN2 rs11079571 could help men figure out how likely they are to get sick.

## Material and methods

### Ethics statement

The KAUST Institutional Review Board in Riyadh, Saudi Arabia, approved the study. All individuals provided written informed consent. Participants were recruited following a physical examination and diagnostic exclusion, in accordance with the IRB’s inclusion criteria.

### Sample collection and Study population

Between XX and XX 2022, individuals meeting the inclusion criteria for medically diagnosed CVD were invited to participate at various locations in Riyadh, Saudi Arabia. Before sample collection, volunteers received informed consent forms, and a team was available to answer any questions about the process. All samples were collected in strict accordance with the ethical standards and human testing methods approved by the KAUST Institutional Review Board (IRB).

For DNA extraction and genotyping, 6 ml of blood samples were collected from each volunteer using K2 EDTA-coated Vacutainer tubes (Cat. no. 23-021-013, K2 EDTA Vacutainer, BD, NJ, USA). The collected samples were transported to the KAUST facility in Riyadh, Saudi Arabia, at 4-8 °C.

A total of 151 blood samples were obtained from individuals with medically diagnosed CVD, aged 52-76 years. Among the samples, 129 were from control individuals who were carefully matched for age. Both the patient and control groups were of Saudi Arabian ethnicity. *Table 1* includes the clinical parameters collected for each individual. The study was powered at 80% to detect an OR ≥ 1.8 for alleles with a frequency of 0.2, ensuring robust statistical findings.

**Table 1.** Clinical parameters of the genotyped participants.

| **Clinical parameter** | **Normal range** | **Male\*** | **Female\*** |
| --- | --- | --- | --- |
| Age (years) | - | 64 ± 12.63 | 64 ± 10.03 |
| Lipid profile (mM) |  |  |  |
| a) Triglycerides | <1.70 | 1.4 | 1.2 |
| b) Total cholesterol | <5.18 | 4.2 | 3.8 |
| c) HDL-cholesterol | >1.04 | 1.0 | 1.2 |
| d) LDL-cholesterol | <2.59 | 2.6 | 2.3 |
| Cardiac enzymes (U/L) |  |  |  |
| a) Aspartate aminotransferase | 6–40 | 69.9 | 20.1 |
| b) Lactate dehydrogenase | 90–200 | 222.8 | 163.8 |
| c) Creatine kinase | 10–150 | 76.7 | 75.3 |

\*Note: Average values listed

### DNA extraction

DNA extraction from blood samples was performed using the QIAamp DNA Blood Mini kit (Cat no: 51106, Qiagen, Valencia, CA, USA). The extraction process for all samples was conducted manually, following the manufacturer’s protocol.

### Quantification and Quality control

Quantification of genomic DNA extracted from blood samples was performed using Qubit® dsDNA BR Assay Kits (Cat no: Q32853, Thermo Fischer Scientific, Massachusetts, USA) on a Qubit reader (Cat no: Q33238, Thermo Fischer Scientific, Massachusetts, USA), following the procedures detailed by the manufacturer. The purity of the samples was assessed using NanoDrop 8000 (Cat no: ND-8000-GL, Thermo Fischer Scientific, Massachusetts, USA) according to the manufacturer’s instructions.

### SNP selection and Genotyping

SNPs from key genes in the WNT signaling pathway were selected from the SNP500 Cancer Project and the relevant literature. We focused on 15 SNPs from 8 genes in the WNT pathway associated with CVD (see *Table 2*). Genotyping was performed using the TaqMan allelic discrimination assay. For PCR reactions, a 5.6 ml 2X universal master mix (Applied Biosystems, Foster City, CA, USA) containing 20 ng DNA and 200 nM primers was prepared. Primers and probe mix were purchased from Applied Biosystems. Genotypes were determined through endpoint reading using an ABI 7500 real-time PCR instrument. To ensure accuracy, 5% of the samples were randomly selected for repeat analysis.

**Table 2.**
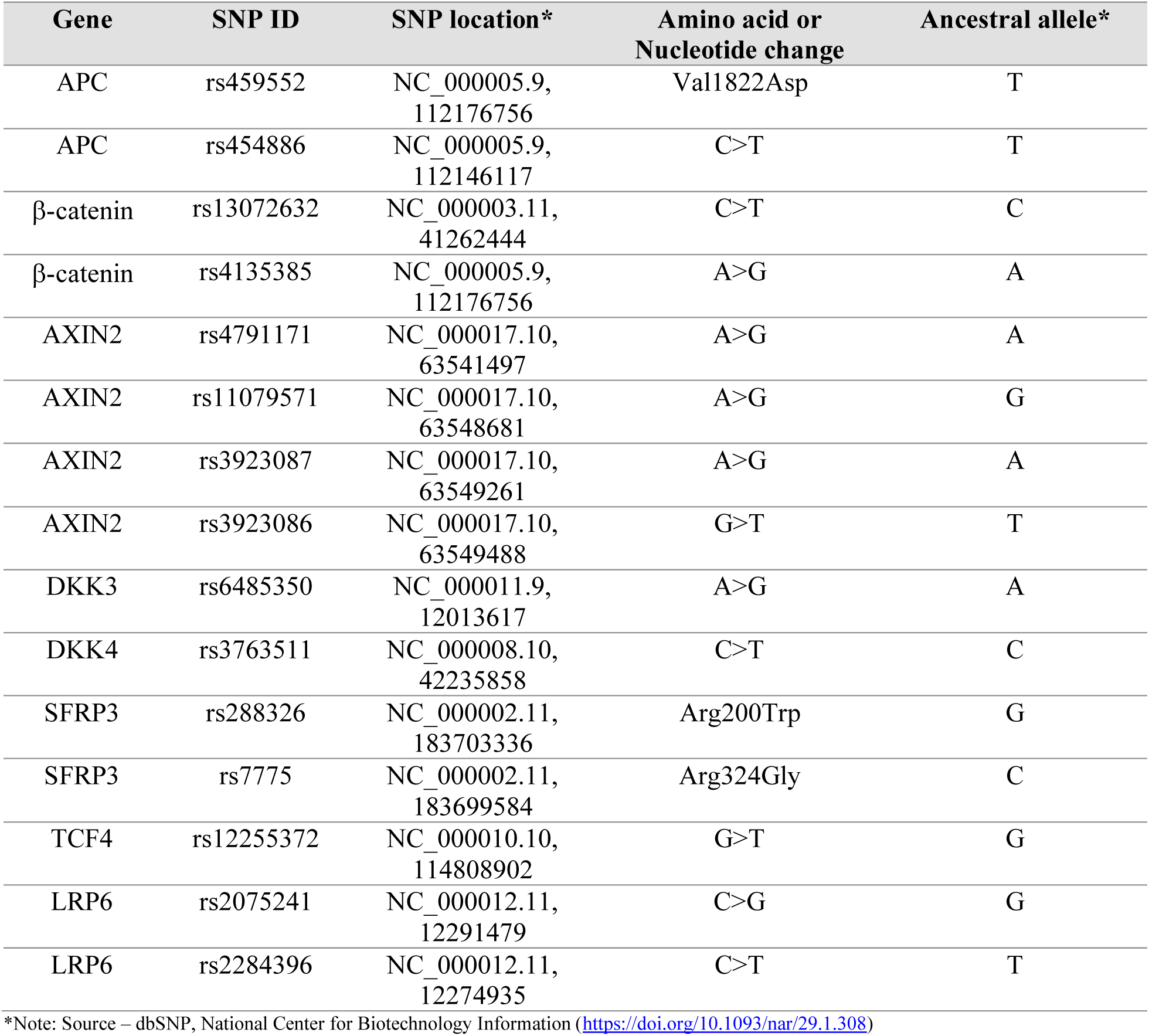
Description of SNPs studied in the current study from WNT pathway genes associated with CVD.

| Gene | SNP ID | SNP location* | Amino acid or Nucleotide change | Ancestral allele* |
| --- | --- | --- | --- | --- |
| APC | rs459552 | NC_000005.9, 112176756 | Val1822Asp | T |
| APC | rs454886 | NC_000005.9, 112146117 | C>T | T |
| β-catenin | rs13072632 | NC_000003.11, 41262444 | C>T | C |
| β-catenin | rs4135385 | NC_000005.9, 112176756 | A>G | A |
| AXIN2 | rs4791171 | NC_000017.10, 63541497 | A>G | A |
| AXIN2 | rs11079571 | NC_000017.10, 63548681 | A>G | G |
| AXIN2 | rs3923087 | NC_000017.10, 63549261 | A>G | A |
| AXIN2 | rs3923086 | NC_000017.10, 63549488 | G>T | T |
| DKK3 | rs6485350 | NC_000011.9, 12013617 | A>G | A |
| DKK4 | rs3763511 | NC_000008.10, 42235858 | C>T | C |
| SFRP3 | rs288326 | NC_000002.11, 183703336 | Arg200Trp | G |
| SFRP3 | rs7775 | NC_000002.11, 183699584 | Arg324Gly | C |
| TCF4 | rs12255372 | NC_000010.10, 114808902 | G>T | G |
| LRP6 | rs2075241 | NC_000012.11, 12291479 | C>G | G |
| LRP6 | rs2284396 | NC_000012.11, 12274935 | C>T | T |

\*Note: Source – dbSNP, National Center for Biotechnology Information (https://doi.org/10.1093/nar/29.1.308)

### Statistical analysis

The risk of disease associated with specific genetic variants was evaluated using odds ratios (ORs). An OR of 1 indicated no association, values less than 1 suggested a protective effect, and values greater than 1 indicated increased risk. The 95% confidence interval (CI) assessed the accuracy and reliability of OR estimates; narrower CIs indicated greater accuracy. Statistical significance and strength of association between each SNP and CVD were determined using P-values: values below 0.05 were considered statistically significant, while higher values suggested a lack of significant association or a possible protective role.

Genotypic and allelic frequencies were calculated for both case and control groups, and conformance to Hardy–Weinberg equilibrium (HWE) was evaluated using a chi-square (χ²) test (http://ihg2.helmholtz-muenchen.de/cgi-bin/hw/hwa1.pl). The goodness-of-fit between observed and expected genotype distributions was assessed independently in cases and controls. Case–control genetic association analyses were performed using Pearson’s chi-square (χ²) test, along with estimation of odds ratios (ORs) and corresponding 95% CIs. Statistical analyses were performed using SPSS software (version XX), and P-values < 0.05 were considered statistically significant.

In addition, the presence and contribution of individual SNPs to CVD susceptibility were further evaluated using association rules based on Certainty Factor, providing an additional measure of the strength and dependability of SNP–disease associations. To assess the overall risk attributable to SNP presence in CVD, a χ² test was used to compare observed and expected distributions, thereby supporting the robustness of the association findings.

## Results

Genetic variation in key Wnt pathway components may influence individual predisposition to CVD. In this study, the effects of Wnt pathway genes on CVD were evaluated by analyzing SNP behavior in 151 patients with CVD and 129 healthy individuals. The analysis focused on fifteen SNPs associated with eight key Wnt genes and explored their roles as protective or risk factors for CVD (*Table 2*). Six SNPs demonstrated both protective and risk-associated roles (*Table 3*). The consequences of these SNPs varied according to age (<58 or >58 years) and gender.

**Table 3.** Genotype frequencies of Wnt pathway gene polymorphism in CVD and Controls.

| Gene | SNP ID | Genotype | CVD<br>(n = ) | Controls<br>(n = ) | OR | (95% CI) | $\chi^2$-Value | P*- Value |
| --- | --- | --- | --- | --- | --- | --- | --- | --- |
| APC | rs459552 | Val | 151 (0.79) | 129 (0.65) | ref |  |  |  |
|  |  | Val/Asp | 37 (0.19) | 64 (0.33) | 0.494 | 0.31-0.79 | 8.88 | 0.00288 |
|  |  | Asp | 3 (0.02) | 4 (0.02) | 0.641 | 0.14-2.92 | 0.34 | 0.56177 |
|  |  | Val/Asp+<br>Asp/Asp | 40 (0.21) | 68 (0.35) | 0.503 | 0.32-0.79 | 8.90 | 0.00286 |
|  |  | Val | 339 (0.89) | 322 (0.82) | ref |  |  |  |
|  |  | Asp | 43 (0.11) | 72 (0.18) | 0.567 | 0.38-0.85 | 7.57 | 0.00595 |
| APC | rs454886 | TT | 27 (0.14) | 22 (0.11) | ref |  |  |  |
|  |  | CT | 69 (0.36) | 80 (0.41) | 0.703 | 0.367-1.344 | 1.14 | 0.28533 |
|  |  | CC | 96 (0.5) | 95 (0.48) | 0.823 | 0.438-1.547 | 0.37 | 0.54537 |
|  |  | CT+CC | 165(0.86) | 175(0.89) | 0.768 | 0.421-1.402 | 0.74 | 0.38960 |
|  |  | T | 123(0.32) | 124(0.31) | ref |  |  |  |
|  |  | C | 261(0.68) | 270(0.69) | 0.975 | 0.721-1.318 | 0.03 | 0.86697 |
| β-Catenin | rs13072632 | CC | 28 (0.14) | 23 (0.12) | ref | | | |
|  |  | CT | 81 (0.43) | 90 (0.46) | 0.739 | 0.39-1.38 | 0.89 | 0.34491 |
|  |  | TT | 81 (0.43) | 84 (0.42) | 0.792 | 0.42-1.49 | 0.53 | 0.46818 |
|  |  | CT+CC | 162 (0.86) | 174 (0.88) | 0.765 | 0.42-1.38 | 0.79 | 0.37337 |
|  |  | C | 137 (0.36) | 136 (0.35) | ref |  |  |  |
|  |  | T | 243 (0.64) | 258 (0.65) | 0.935 | 0.7-1.26 | 0.20 | 0.65504 |
| β-Catenin | rs4135385 | AA | 146 (0.76) | 143 (0.73) | ref | | | |
|  |  | AG | 40 (0.21) | 47 (0.23) | 0.834 | 0.52-1.35 | 0.55 | 0.45755 |
|  |  | GG | 5 (0.03) | 7 (0.04) | 0.700 | 0.22-2.25 | 0.36 | 0.54786 |
|  |  | AG+GG | 45 (0.24) | 54 (0.27) | 0.816 | 0.52-1.29 | 0.76 | 0.38436 |
|  |  | A | 332 (0.87) | 333 (0.85) | ref |  |  |  |
|  |  | G | 50 (0.13) | 61 (0.15) | 0.822 | 0.55-1.23 | 0.91 | 0.34111 |
| AXIN2 | rs4791171 | AA | 56 (0.30) | 68 (0.35) | ref |  |  |  |
|  |  | AG | 81 (0.43) | 89 (0.45) | 1.105 | 0.69-1.76 | 0.18 | 0.67306 |
|  |  | GG | 51 (0.27) | 40 (0.2) | 1.548 | 0.9-2.67 | 2.49 | 0.11484 |
|  |  | AG+GG | 132 (0.7) | 129 (0.65) | 1.243 | 0.81-1.91 | 0.99 | 0.32074 |
|  |  | A | 193 (0.51) | 225 (0.57) | ref |  |  |  |
|  |  | G | 183 (0.49) | 169 (0.43) | 1.262 | 0.95-1.68 | 2.59 | 0.10773 |
| AXIN2 | rs11079571 | GG | 75 (0.40) | 88(0.47) | Ref |  |  |  |
|  |  | GA | 78 (0.42) | 69(0.37) | 1.326 | 0.848-2.074 | 1.54 | 0.21514 |
|  |  | AA | 34 (0.18) | 32(0.16) | 1.247 | 0.703-2.210 | 0.57 | 0.45012 |
|  |  | GA+AA | 112(0.60) | 101(0.53) | 1.301 | 0.864-1.958 | 1.59 | 0.20671 |
|  |  | G | 228(0.61) | 245(0.65) | Ref |  |  |  |
|  |  | A | 146(0.39) | 133(0.35) | 1.180 | 0.877-1.586 | 1.20 | 0.27423 |
| **AXIN2** | rs3923087 | AA | 51 (0.31) | 60 (0.3) | ref |  |  |  |
|  |  | AG | 86 (0.53) | 95 (0.48) | 1.065 | 0.66-1.71 | 0.07 | 0.79440 |
|  |  | GG | 54 (0.16) | 42 (0.22) | 0.728 | 0.39-1.35 | 1.02 | 0.31186 |
|  |  | AG+GG | 140 (0.69) | 137 (0.7) | 0.962 | 0.61-1.51 | 0.03 | 0.86497 |
|  |  | A | 188 (0.58) | 215 (0.55) | ref |  |  |  |
|  |  | G | 194 (0.42) | 179 (0.45) | 0.882 | 0.66-1.18 | 0.70 | 0.40421 |
| **AXIN2** | rs3923086 | TT | 24 (0.13) | 35 (0.18) | ref |  |  |  |
|  |  | TG | 90 (0.47) | 89 (0.45) | 1.475 | 0.812-2.677 | 1.64 | 0.20044 |
|  |  | GG | 78 (0.40) | 73 (0.37) | 1.558 | 0.847-2.867 | 2.05 | 0.15254 |
|  |  | TG+GG | 168(0.87) | 162(0.82) | 1.512 | 0.862-2.654 | 2.10 | 0.14768 |
|  |  | T | 138(0.36) | 159(0.40) | ref |  |  |  |
|  |  | G | 246(0.64) | 235(0.60) | 1.206 | 0.903-1.611 | 1.61 | 0.20475 |
| **DKK3** | rs6485350 | AA | 63 (0.33) | 56 (0.31) | ref |  |  |  |
|  |  | AG | 98 (0.51) | 85 (0.46) | 1.025 | 0.64-1.63 | 0.01 | 0.91721 |
|  |  | GG | 30 (0.16) | 42 (0.23) | 0.635 | 0.35-1.15 | 2.28 | 0.13085 |
|  |  | AG+GG | 128 (0.67) | 127 (0.69) | 0.896 | 0.58-1.38 | 0.24 | 0.62085 |
|  |  | A | 224 (0.59) | 197 (0.54) | ref |  |  |  |
|  |  | G | 158 (0.41) | 169 (0.46) | 0.822 | 0.62-1.1 | 1.76 | 0.18460 |
| **DKK4** | rs3763511 | CC | 130 (0.68) | 148 (0.75) | ref |  |  |  |
|  |  | CT | 52 (0.27) | 48 (0.24) | 1.233 | 0.78-1.95 | 0.81 | 0.36870 |
|  |  | TT | 10 (0.05) | 1 (0.01) | 11.385 | 1.44-90.14 | 8.26 | 0.00406 |
|  |  | CT+CC | 62 (0.32) | 49 (0.25) | 1.441 | 0.92-2.24 | 2.62 | 0.10525 |
|  |  | C | 312 (0.81) | 344 (0.87) | ref |  |  |  |
|  |  | T | 72 (0.19) | 50 (0.13) | 1.588 | 1.07-2.35 | 5.40 | 0.02013 |
| **SFRP3** | rs288326 | AA | 1 (0.01) | 4 (0.02) | ref |  |  |  |
|  |  | AG | 28 (0.14) | 32 (0.16) | 3.500 | 0.37-33.18 | 1.33 | 0.24912 |
|  |  | GG | 161 (0.85) | 160 (0.82) | 4.025 | 0.44-36.41 | 1.79 | 0.18081 |
|  |  | AG+GG | 189 (0.99) | 192 (0.98) | 3.938 | 0.44-35.55 | 1.73 | 0.18831 |
|  |  | A | 30 (0.08) | 40 (0.10) | ref |  |  |  |
|  |  | G | 350 (0.92) | 352 (0.9) | 1.326 | 0.81-2.18 | 1.25 | 0.26393 |
| **SFRP3** | rs7775 | Arg | 3 (0.02) | 4 (0.02) | ref |  |  |  |
|  |  | Arg/Gly | 40 (0.21) | 55 (0.28) | 0.970 | 0.206-4.575 | 0 | 0.96899 |
|  |  | Gly | 148 (0.77) | 138 (0.7) | 1.430 | 0.314-6.504 | 0.22 | 0.64190 |
|  |  | Arg/Gly+Gly | 188 (0.98) | 193 (0.98) | 1.299 | 0.287-5.881 | 0.12 | 0.73373 |
|  |  | Arg | 46 (0.12) | 63 (0.16) | ref |  |  |  |
|  |  | Gly | 302 (0.88) | 331 (0.84) | 1.390 | 0.923-2.093 | 2.50 | 0.11357 |
| **TCF7L2** | rs12255372 | GG | 80 (0.41) | 73 (0.37) | ref |  |  |  |
|  |  | GT | 86 (0.45) | 94 (0.48) | 0.835 | 0.542-1.285 | 0.67 | 0.41207 |
|  |  | TT | 26 (0.14) | 30 (0.15) | 0.791 | 0.428-1.461 | 0.56 | 0.45304 |
|  |  | GT+TT | 112 (0.59) | 124 (0.63) | 0.824 | 0.548-1.239 | 0.87 | 0.35198 |
|  |  | G | 246 (0.64) | 240 (0.61) | ref |  |  |  |
|  |  | T | 138 (0.36) | 154 (0.39) | 0.874 | 0.654-1.169 | 0.82 | 0.36448 |
| **LRP6** | rs2075241 | GG | 129 (0.68) | 124 (0.64) | ref |  |  |  |
|  |  | GC | 57 (0.3) | 64 (0.33) | 0.856 | 0.555-1.321 | 0.49 | 0.48255 |
|  |  | CC | 5 (0.02) | 7 (0.03) | 0.687 | 0.212-2.221 | 0.40 | 0.52800 |
|  |  | GC+CC | 62(0.32) | 71(0.36) | 0.839 | 0.551-1.278 | 0.67 | 0.41429 |
|  |  | G | 315(0.82) | 312(0.80) | ref |  |  |  |
|  |  | C | 67(0.18) | 78(0.20) | 0.851 | 0.592-1.222 | 0.77 | 0.38145 |
| **LRP6** | rs2284396 | TT | 90 (0.47) | 106 (0.54) | ref |  |  |  |
|  |  | TC | 90 (0.47) | 69 (0.35) | 1.536 | 1.008-2.341 | 4.01 | 0.04523 |
|  |  | CC | 12 (0.06) | 21 (0.11) | 0.673 | 0.314-1.443 | 1.04 | 0.30693 |
|  |  | TC+CC | 102(0.53) | 90(0.46) | 1.335 | 0.896-1.989 | 2.02 | 0.15574 |
|  |  | T | 270(0.72) | 281(0.70) | ref |  |  |  |
|  |  | C | 114(0.28) | 111(0.30) | 1.069 | 0.784-1.458 | 0.18 | 0.67384 |

Note: OR 95% CI. Odds Ratio and 95% Confidence Interval. \* P < 0.05 was considered significant

### 1. APC gene

In the present study, two APC-associated SNPs—rs459552 (Val/Asp) and rs454886—showed a significant association with CVD risk, with both variants exhibiting protective effects (*Table 3*). The SNP rs459552 showed a protective association in the heterozygous Val/Asp genotype (OR = 0.494, p = 0.003), while rs454886 was associated with reduced CVD risk in the homozygous CC genotype, particularly among male participants (OR = 0.335, p = 0.008). This protective association was gender-and age-dependent, being significant in males but not females (*Tables 4 and 5*), and observed predominantly in individuals below 58 years of age, suggesting an age-related attenuation of genetic protection (*Tables 6 and 7*). Specifically, rs459552 showed a robust protective association in patients younger than 58 years (OR = 0.468, p = 0.02).

**Table 4.**
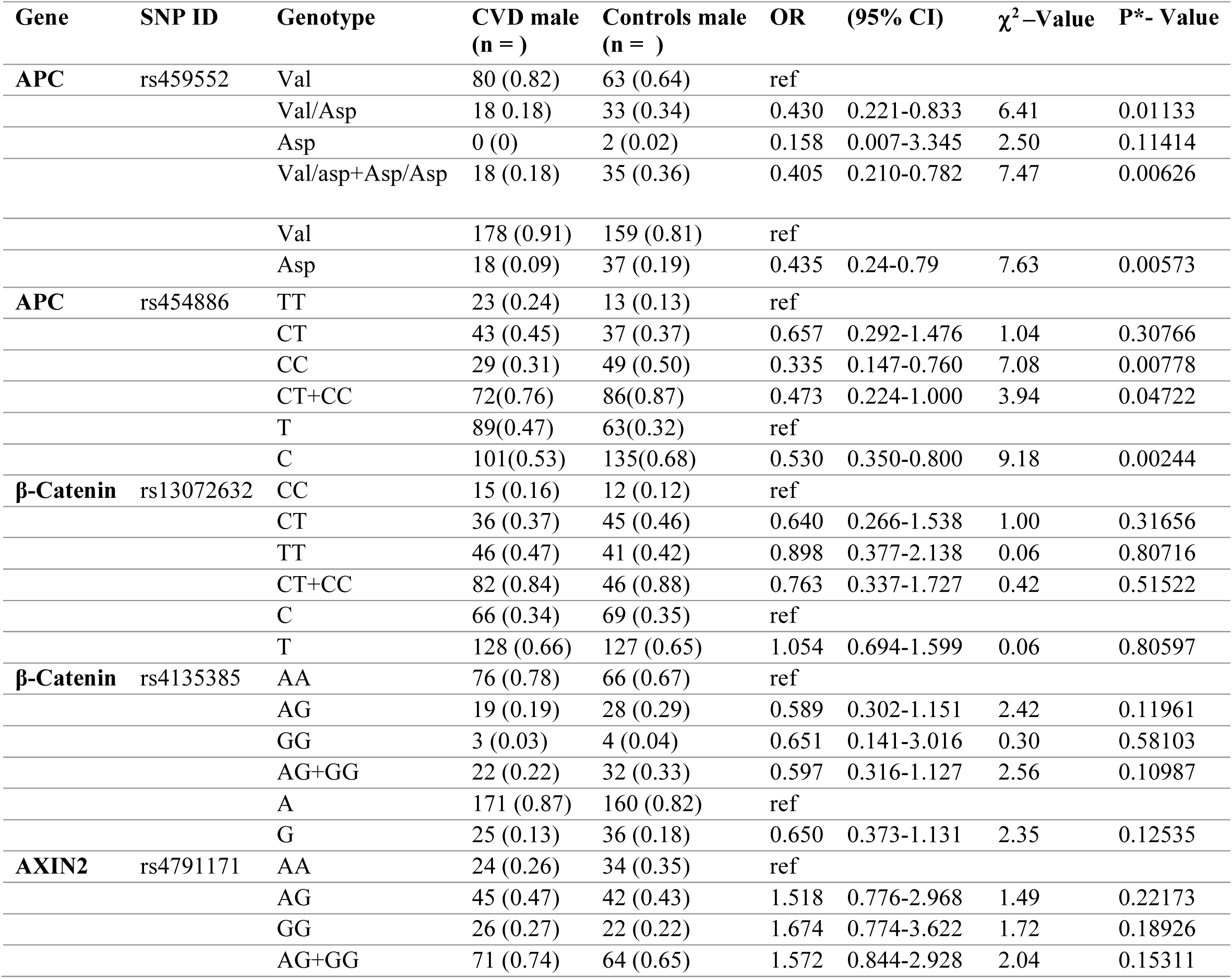
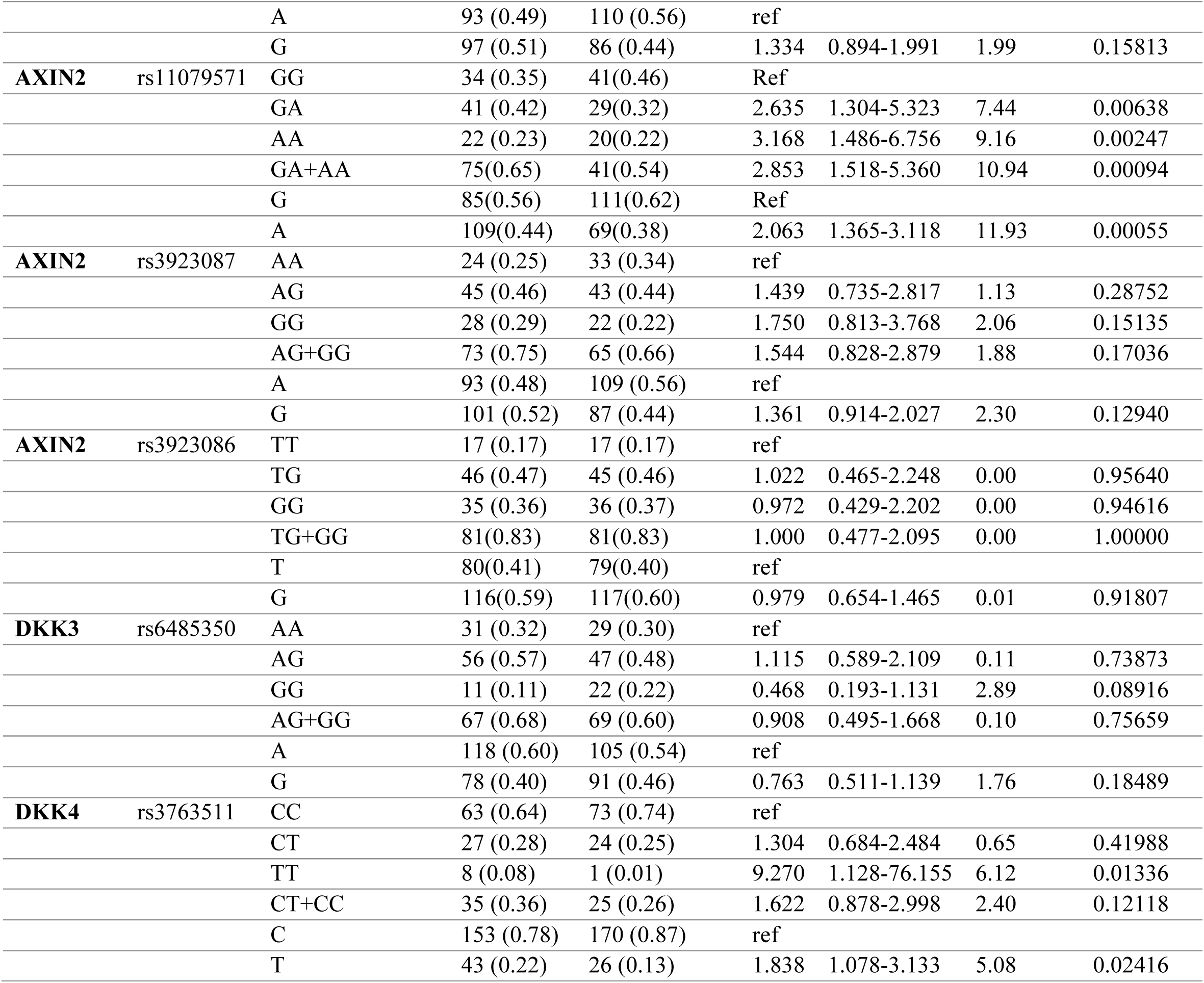
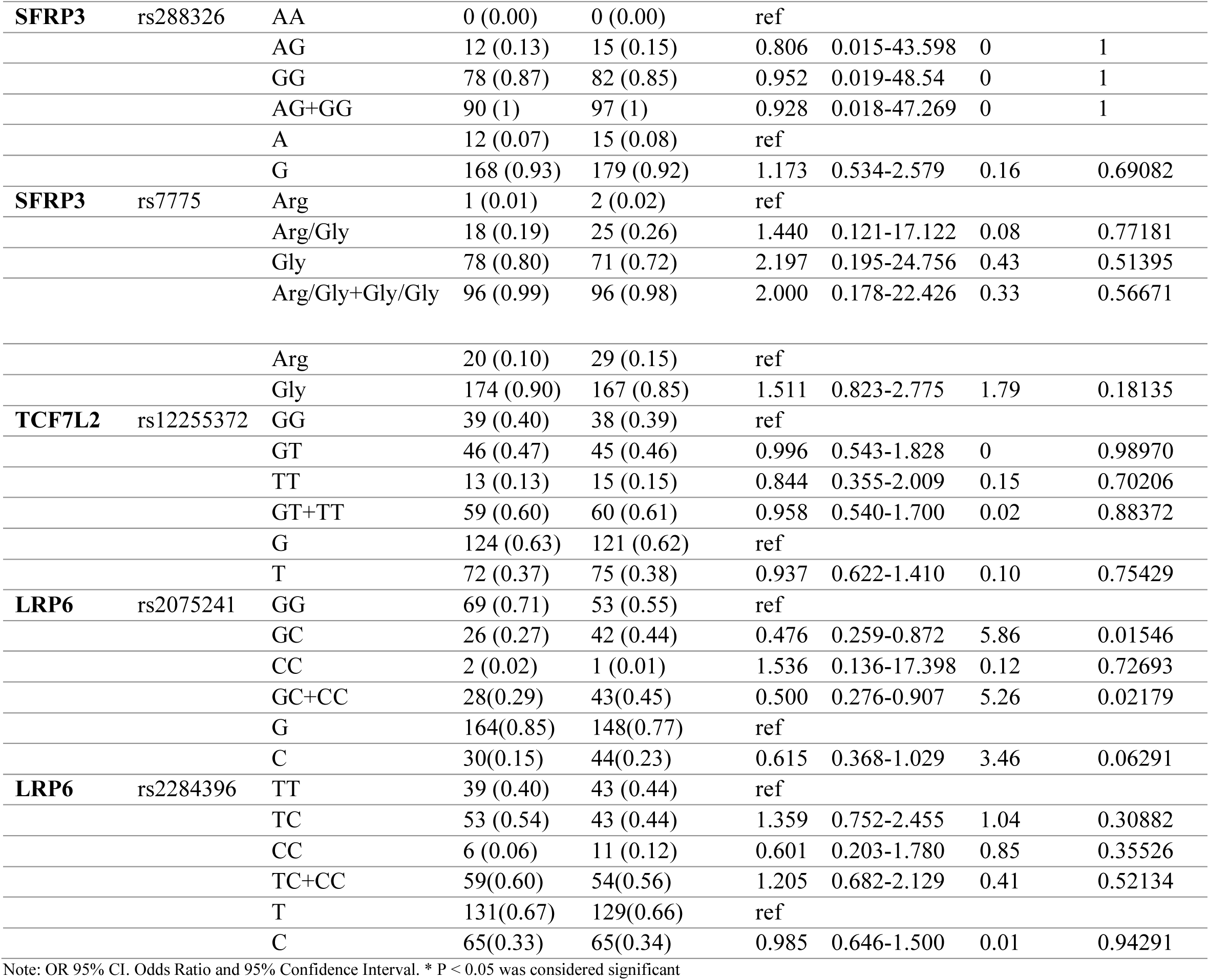
Genotype Frequencies of Wnt gene Polymorphism in between CVD male Cases and controls.

**Table 5.**
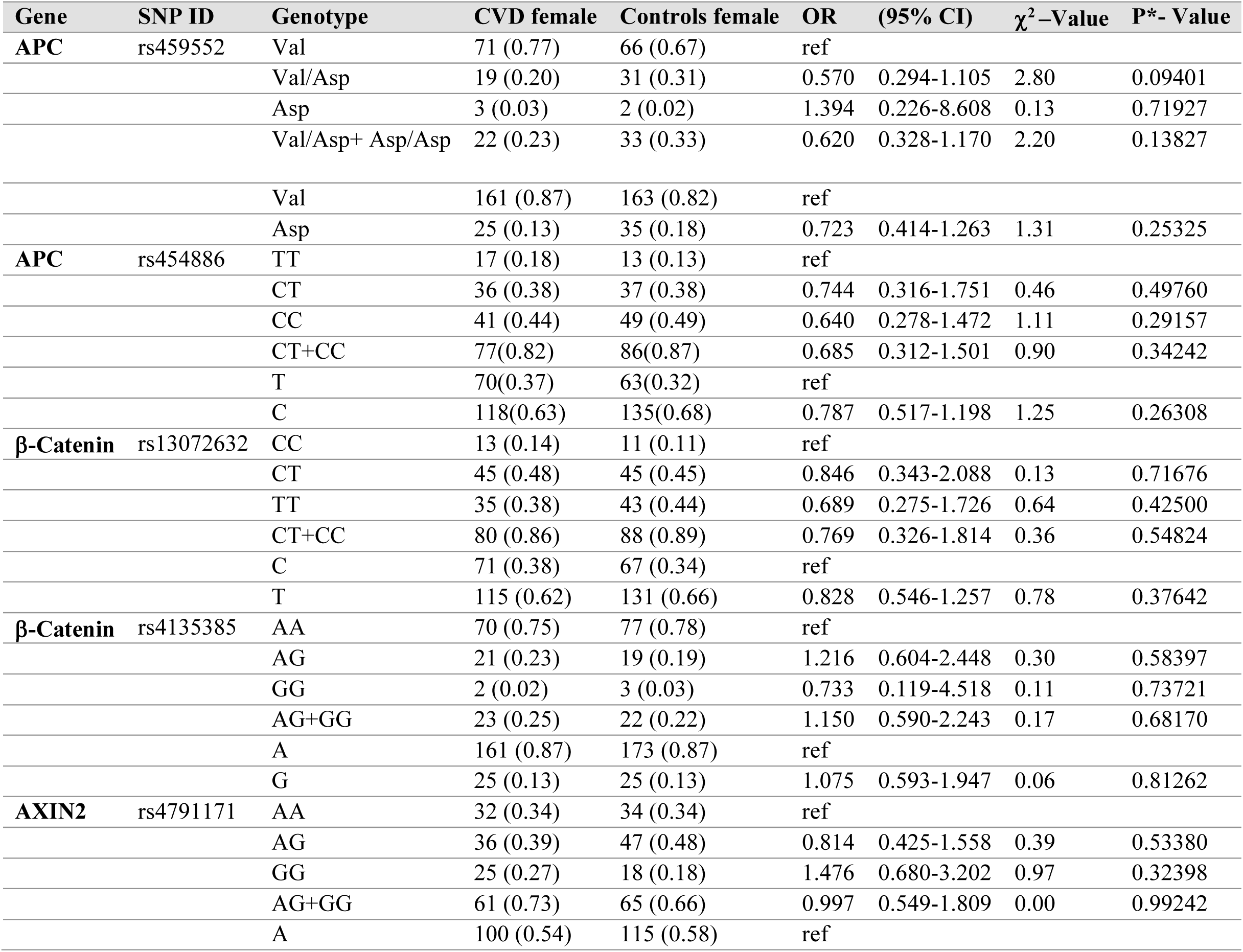
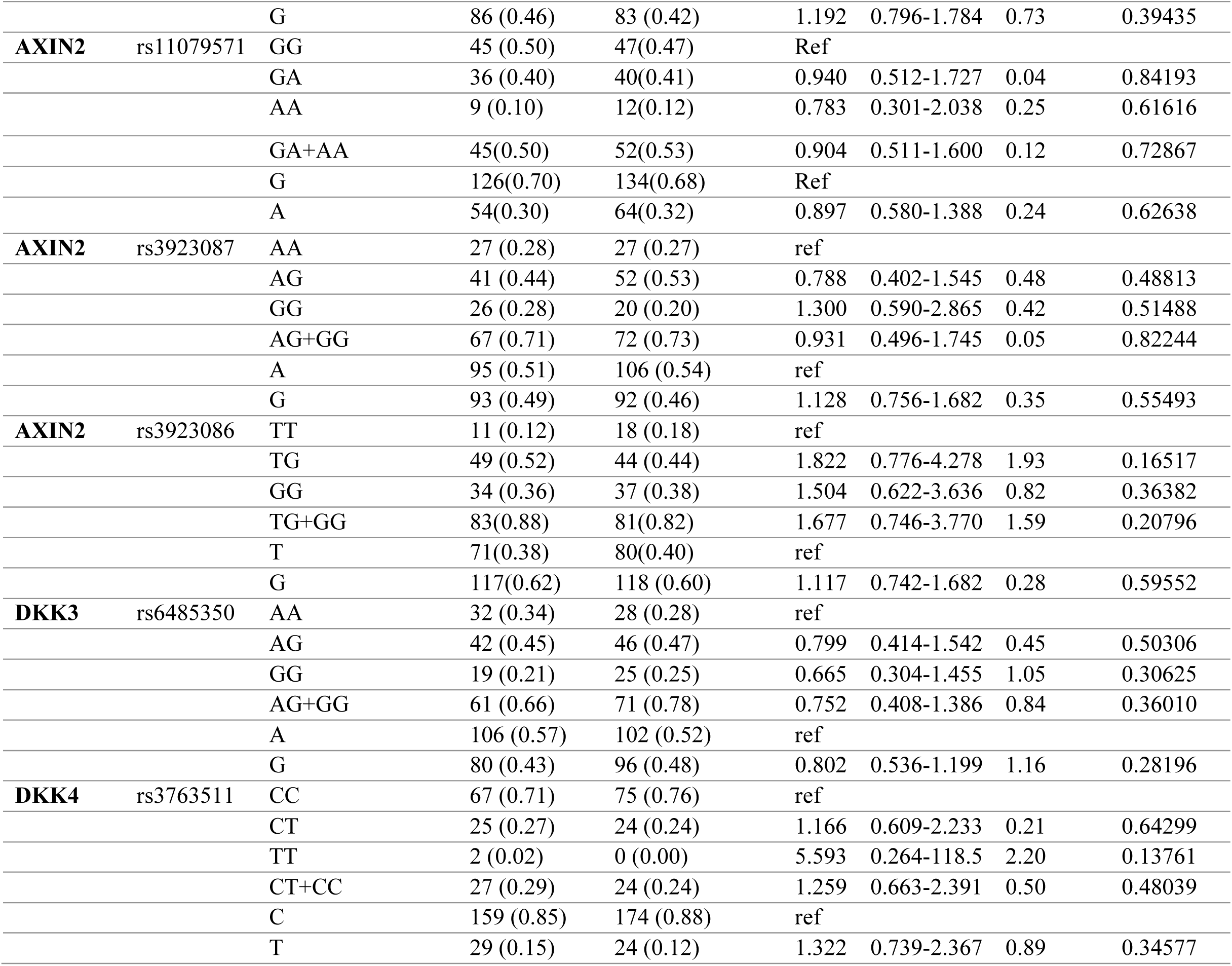
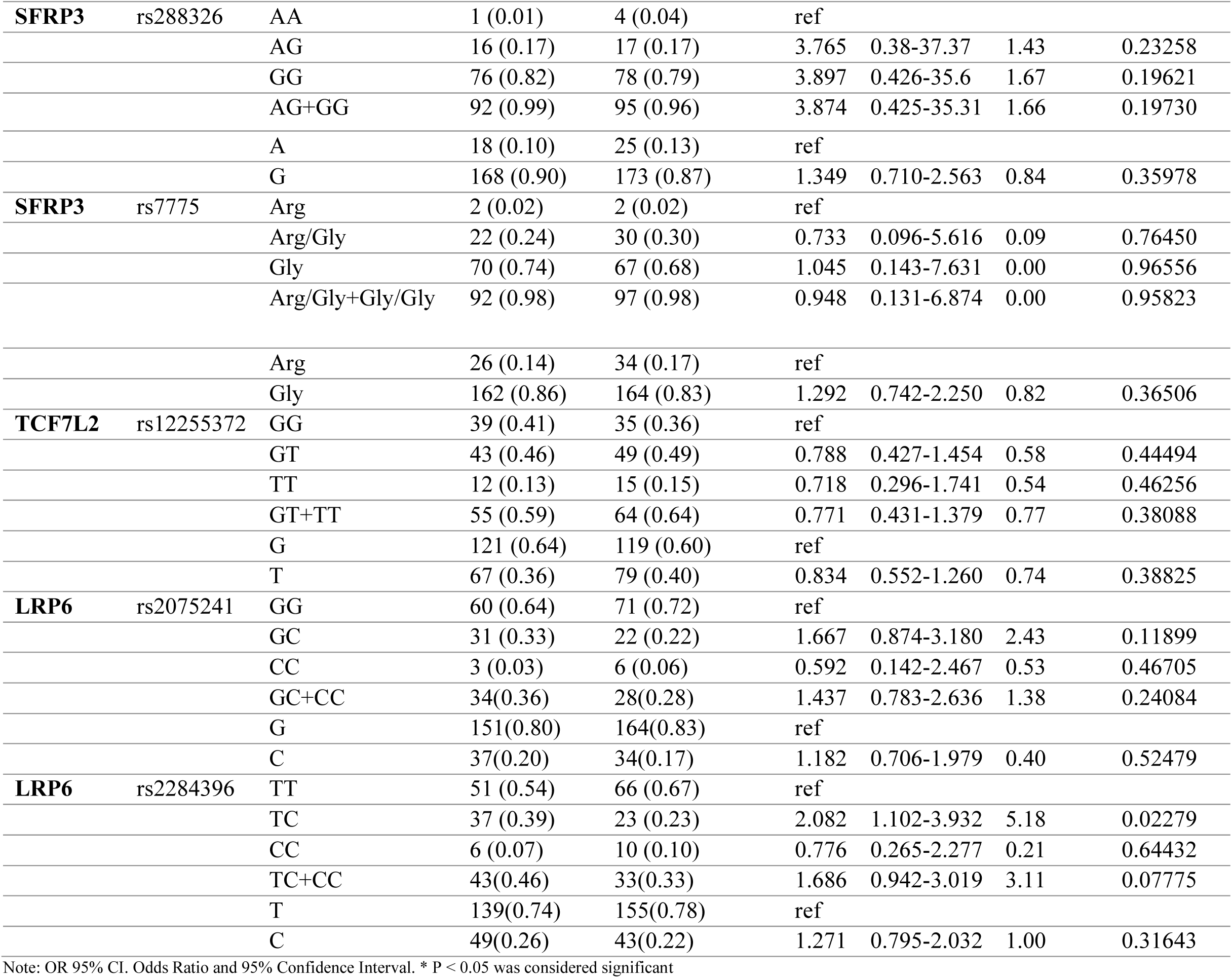
Genotype Frequencies of wnt Gene Polymorphism in between CVD female Cases and controls.

**Table 6.** Genotype Frequencies of Wnt gene Polymorphism in between CVD below 58y aged Cases and controls.

| Gene | SNP ID | Genotype | CVD male | Controls male | OR | (95% CI) | $\chi^2$-Value | P*- Value |
| --- | --- | --- | --- | --- | --- | --- | --- | --- |
| APC | rs459552 | Val | 69 (0.78) | 68(0.62) | ref |  |  |  |
|  |  | Val/ Asp | 19 (0.21) | 40(0.36) | 0.468 | 0.247-0.888 | 5.50 | 0.01903 |
|  |  | Asp | 1 (0.01) | 2(0.02) | 0.493 | 0.044-5.562 | 0.34 | 0.55946 |
|  |  | Val/asp+Asp/ Asp | 20 (0.22) | 42 (0.38) | 0.469 | 0.250-0.880 | 5.66 | 0.01735 |
|  |  | Val | (0.88) | (0.80) | ref |  |  |  |
|  |  | Asp | (0.12) | (0.20) | 0.535 | 0.305-0.939 | 4.84 | 0.02774 |
| APC | rs454886 | TT | 14(0.16) | 14(0.13) | ref |  |  |  |
|  |  | CT | 41(0.47) | 42(0.38) | 0.976 | 0.414-2.299 | 0.00 | 0.95603 |
|  |  | CC | **33(0.37)** | 54(0.49) | 0.611 | 0.259-1.441 | 1.28 | 0.25850 |
|  |  | CT+CC | **74(0.84)** | 96(0.87) | 0.771 | 0.346-1.716 | 0.41 | 0.52317 |
|  |  | T | 69(0.39) | 70(0.32) | ref |  |  |  |
|  |  | C | 107(0.61) | 150(0.68) | 0.724 | 0.478-1.096 | 2.34 | 0.12595 |
| β-Catenin | rs13072632 | CC | 13(0.15) | 10(0.09) | ref | | | |
|  |  | CT | 34(0.38) | 48(0.44) | 0.545 | 0.214-1.387 | 1.65 | 0.19933 |
|  |  | TT | 42(0.47) | 52(0.47) | 0.621 | 0.248-1.558 | 1.04 | 0.30781 |
|  |  | CT+CC | 76 (0.85) | 100 (0.91) | 0.585 | 0.243-1.405 | 1.46 | 0.22625 |
|  |  | C | 60(0.34) | 68(0.31) | ref |  |  |  |
|  |  | T | 118(0.66) | 152(0.69) | 0.880 | 0.577-1.342 | 0.35 | 0.55228 |
| β-Catenin | rs4135385 | AA | 68(0.77) | 80(0.73) | ref | | | |
|  |  | AG | 18(0.20) | 24(0.22) | 0.882 | 0.442-1.762 | 0.13 | 0.72263 |
|  |  | GG | 3(0.03) | 6(0.05) | 0.588 | 0.142-2.441 | 0.54 | 0.46044 |
|  |  | AG+GG | 21 (0.23) | 30 (0.27) | 0.824 | 0.432-1.569 | 0.35 | 0.55467 |
|  |  | A | 154(0.87) | 184(0.84) | ref |  |  |  |
|  |  | G | 24(0.13) | 36(0.16) | 0.797 | 0.455-1.393 | 0.64 | 0.42456 |
| AXIN2 | rs4791171 | AA | 27(0.31) | 35(0.32) | ref |  |  |  |
|  |  | AG | 35(0.39) | 53(0.48) | 0.856 | 0.443-1.655 | 0.21 | 0.64378 |
|  |  | GG | 26(0.30) | 22(0.20) | 1.532 | 0.718-3.269 | 1.22 | 0.26902 |
|  |  | AG+GG | 61(0.69) | 75 (0.68) | 1.054 | 0.576-1.931 | 0.03 | 0.86397 |
|  |  | A | 89(0.51) | 123(0.56) | ref |  |  |  |
|  |  | G | 87(0.49) | 97(0.44) | 1.240 | 0.833-1.845 | 1.12 | 0.28965 |
| **AXIN2** | rs11079571 | GG | 35(0.39) | 58(0.53) | ref |  |  |  |
|  |  | GA | 35(0.39) | 33(0.30) | 1.758 | 0.932-3.314 | 3.06 | 0.08025 |
|  |  | AA | 20(0.22) | 19(0.17) | 1.744 | 0.820-3.712 | 2.11 | 0.14676 |
|  |  | GA+AA | 55(0.61) | 52(0.47) | 1.753 | 0.996-3.085 | 3.81 | 3.81 |
|  |  | G | 105(0.58) | 149(0.68) | ref |  |  |  |
|  |  | A | 75(0.42) | 71(0.32) | 1.499 | 0.995-2.258 | 3.77 | 0.05220 |
| **AXIN2** | rs3923087 | AA | 25(0.28) | 30(0.27) | ref |  |  |  |
|  |  | AG | 39(0.43) | 55(0.50) | 0.851 | 0.435-1.664 | 0.22 | 0.63704 |
|  |  | GG | 26(0.29) | 25(0.23) | 1.248 | 0.582-2.678 | 0.32 | 0.56941 |
|  |  | AG+GG | 65 (0.72) | 80 (0.73) | 0.975 | 0.523-1.819 | 0.01 | 0.93657 |
|  |  | A | 89(0.49) | 115(0.52) | ref |  |  |  |
|  |  | G | 91(0.51) | 105(0.48) | 1.120 | 0.755-1.661 | 0.32 | 0.57348 |
| **AXIN2** | rs3923086 | TT | 16(0.18) | 16(0.15) | ref |  |  |  |
|  |  | TG | 45(0.50) | 55(0.50) | 0.818 | 0.369-1.816 | 0.24 | 0.62146 |
|  |  | GG | 29(0.32) | 39(0.35) | 0.744 | 0.320-1.728 | 0.48 | 0.49054 |
|  |  | TG+GG | 74(0.82) | 94(0.85) | 0.787 | 0.369-1.678 | 0.38 | 0.53505 |
|  |  | T | 77(0.43) | 87(0.40) | ref |  |  |  |
|  |  | G | 103(0.57) | 133(0.60) | 0.875 | 0.586-1.306 | 0.43 | 0.51317 |
| **DKK3** | rs6485350 | AA | 31(0.35) | 29(0.26) | ref |  |  |  |
|  |  | AG | 43(0.48) | 53(0.48) | 0.759 | 0.398-1.449 | 0.70 | 0.40281 |
|  |  | GG | 15(0.17) | 28(0.26) | 0.501 | 0.224-1.122 | 2.85 | 0.09111 |
|  |  | AG+GG | 58 (0.65) | 81 (0.74) | 0.670 | 0.365-1.231 | 1.67 | 0.19559 |
|  |  | A | 105(0.59) | 111(0.50) | ref |  |  |  |
|  |  | G | 73(0.41) | 109(0.50) | 0.708 | 0.475-1.055 | 2.89 | 0.08927 |
| **DKK4** | rs3763511 | CC | 62(0.69) | 84(0.76) | ref |  |  |  |
|  |  | CT | 25(0.28) | 25(0.23) | 1.355 | 0.711-2.580 | 0.86 | 0.35473 |
|  |  | TT | 3(0.03) | 1(0.01) | 4.065 | 0.413-40.010 | 1.68 | 0.19516 |
|  |  | CT+CC | 28 (0.31) | 26 (0.24) | 1.459 | 0.780-2.730 | 1.40 | 0.23619 |
|  |  | C | 149(0.83) | 193(0.88) | ref |  |  |  |
|  |  | T | 31(0.17) | 27(0.12) | 1.487 | 0.851-2.600 | 1.96 | 0.16192 |
| **SFRP3** | rs288326 | AA | 0(0.00) | 2(0.02) | ref |  |  |  |
|  |  | AG | 14(0.16) | 18(0.16) | 3.919 | 0.174-88.149 | 1.49 | 0.22260 |
|  |  | GG | 75(0.84) | 90(0.82) | 4.171 | 0.197-88.227 | 1.65 | 0.19893 |
|  |  | AG+GG | 89 (1) | 108 (0.98) | 4.124 | 0.195-87.022 | 1.63 | 0.20107 |
|  |  | A | (0.08) | (0.10) | ref |  |  |  |
|  |  | G | (0.92) | (0.90) | 1.302 | 0.646-2.624 | 0.55 | 0.46034 |
| **SFRP3** | rs7775 | Arg | 1(0.01) | 2(0.02) | ref |  |  |  |
|  |  | Arg/Gly | 20(0.22) | 25(0.23) | 1.600 | 0.135-18.943 | 0.14 | 0.70720 |
|  |  | Gly | 68(0.77) | 83(0.75) | 1.639 | 0.145-18.460 | 0.16 | 0.68657 |
|  |  | Arg/Gly+Gly/Gly | 88 (0.99) | 108 (0.98) | 1.630 | 0.145-18.270 | 0.16 | 0.68930 |
|  |  | Arg | 22(0.12) | 29(0.13) | ref |  |  |  |
|  |  | Gly | 156(0.88) | 191(0.87) | 1.077 | 0.595-1.948 | 0.06 | 0.80721 |
| **TCF7L2** | rs12255372 | GG | 38(0.42) | 39(0.36) | ref |  |  |  |
|  |  | GT | 41(0.46) | 53(0.48) | 0.794 | 0.434-1.454 | 0.56 | 0.45433 |
|  |  | TT | 11(0.12) | 18(0.16) | 0.627 | 0.262-1.502 | 1.11 | 0.29314 |
|  |  | GT+TT | 62 (0.58) | 71 (0.64) | 0.752 | 0.424-1.332 | 0.96 | 0.32781 |
|  |  | G | 117(0.65) | 131(0.60) | ref |  |  |  |
|  |  | T | 63(0.35) | 89(0.40) | 0.793 | 0.527-1.192 | 1.25 | 0.26352 |
| **LRP6** | rs2075241 | GG | 56(0.63) | 67(0.62) | ref |  |  |  |
|  |  | GC | 28(0.31) | 37(0.34) | 0.905 | 0.494-1.659 | 0.10 | 0.74778 |
|  |  | CC | 5(0.06) | 4(0.04) | 1.496 | 0.383-5.838 | 0.34 | 0.56028 |
|  |  | GC+CC | 33 (0.37) | 41 (0.38) | 0.963 | 0.539-1.719 | 0.02 | 0.89851 |
|  |  | G | 140 (0.79) | 171 (0.79) | ref |  |  |  |
|  |  | C | 38 (0.21) | 45 (0.21) | 1.031 | 0.634-1.677 | 0.02 | 0.90072 |
| **LRP6** | rs2284396 | TT | 49(0.54) | 67(0.62) | ref |  |  |  |
|  |  | TC | 33(0.37) | 33(0.30) | 1.367 | 0.745-2.509 | 1.02 | 0.31183 |
|  |  | CC | 8(0.09) | 9(0.08) | 1.215 | 0.438-3.374 | 0.14 | 0.70778 |
|  |  | TC+CC | 41 (0.46) | 42 (0.38) | 1.335 | 0.758-2.352 | 1.00 | 0.31726 |
|  |  | T | 131 (0.67) | 167 (0.77) | ref |  |  |  |
|  |  | C | 49 (0.33) | 51 (0.23) | 1.225 | 0.778-1.928 | 0.77 | 0.38088 |

Note: OR 95% CI. Odds Ratio and 95% Confidence Interval. \* P < 0.05 was considered significant.

**Table 7.** Genotype Frequencies of Wnt gene Polymorphism in between CVD above 58y aged Cases and controls.

| Gene | SNP ID | Genotype | CVD male | Controls male | OR | (95% CI) | $\chi^2$-Value | P*- Value |
| --- | --- | --- | --- | --- | --- | --- | --- | --- |
| APC | rs459552 | Val | 76(0.81) | 61(0.70) | ref |  |  |  |
|  |  | Val/Asp | 17(0.18) | 24(0.28) | 0.569 | 0.280-1.153 | 2.48 | 0.11509 |
|  |  | Asp | 1(0.01) | 2(0.02) | 0.401 | 0.036-4.531 | 0.58 | 0.44573 |
|  |  | Val/asp+Asp/Asp | 18 (0.19) | 26 (0.30) | 0.556 | 0.279-1.107 | 2.83 | 0.09250 |
|  |  | Val | 169(0.90) | 146(0.84) | ref |  |  |  |
|  |  | Asp | 19(0.10) | 28(0.16) | 0.586 | 0.314-1.093 | 2.87 | 0.09049 |
| APC | rs454886 | TT | 23(0.24) | 8(0.09) | ref |  |  |  |
|  |  | CT | 36(0.39) | 38(0.44) | 0.330 | 0.131-0.831 | 5.79 | 0.01611 |
|  |  | CC | 34(0.37) | 41(0.47) | 0.288 | 0.114-0.727 | 7.35 | 0.00671 |
|  |  | CT+CC | 70 (0.76) | 79 (0.91) | 0.308 | 0.130-0.733 | 7.61 | 0.00580 |
|  |  | T | 82 (0.44) | 54 (0.31) | ref |  |  |  |
|  |  | C | 104 (0.56) | 120(0.69) | 0.571 | 0.370-0.879 | 6.51 | 0.01070 |
| β-Catenin | rs13072632 | CC | 13(0.14) | 13(0.15) | ref | | | |
|  |  | CT | 45(0.48) | 42(0.48) | 1.071 | 0.446-2.573 | 0.02 | 0.87735 |
|  |  | TT | 35(0.38) | 32(0.37) | 1.094 | 0.442-2.706 | 0.04 | 0.84626 |
|  |  | CT+CC | 80 (0.86) | 74 (0.85) | 1.081 | 0.471-2.482 | 0.03 | 0.85412 |
|  |  | C | 71(0.38) | 68(0.39) | ref |  |  |  |
|  |  | T | 115(0.62) | 106(0.61) | 1.039 | 0.680-1.589 | 0.03 | 0.85958 |
| β-Catenin | rs4135385 | AA | 71(0.76) | 63(0.73) | ref | | | |
|  |  | AG | 21(0.22) | 23(0.26) | 0.810 | 0.410-1.602 | 0.37 | 0.54481 |
|  |  | GG | 2(0.02) | 1(0.01) | 1.775 | 0.157-20.044 | 0.22 | 0.63854 |
|  |  | AG+GG | 23 (0.24) | 24 (0.27) | 0.850 | 0.437-1.653 | 0.23 | 0.63263 |
|  |  | A | 163(0.87) | 149(0.86) | ref |  |  |  |
|  |  | G | 25(0.13) | 25(0.14) | 0.914 | 0.503-1.661 | 0.09 | 0.76816 |
| AXIN2 | rs4791171 | AA | 23(0.25) | 33(0.38) | ref |  |  |  |
|  |  | AG | 45(0.49) | 36(0.41) | 1.793 | 0.900-3.574 | 2.78 | 0.09553 |
|  |  | GG | 24(0.26) | 18(0.21) | 1.913 | 0.851-4.302 | 2.48 | 0.11503 |
|  |  | AG+GG | 69 (0.75) | 54 (0.62) | 1.833 | 0.966-3.479 | 3.48 | 0.06219 |
|  |  | A | 91(0.49) | 102(0.59) | ref |  |  |  |
|  |  | G | 93(0.51) | 72(0.41) | 1.448 | 0.953-2.199 | 3.02 | 0.08211 |
| **AXIN2** | rs11079571 | GG | 32(0.36) | 30(0.38) | ref |  |  |  |
|  |  | GA | 38(0.43) | 36(0.46) | 0.990 | 0.504-1.944 | 0.00 | 0.97575 |
|  |  | AA | 19(0.21) | 13(0.16) | 1.370 | 0.578-3.250 | 0.51 | 0.47411 |
|  |  | GA+AA | 57 (0.66) | 49(0.62) | 1.091 | 0.582-2.043 | 0.07 | 0.78658 |
|  |  | G | 102 (0.57) | 96(0.61) | ref |  |  |  |
|  |  | A | 106 (0.43) | 62(0.39) | 1.154 | 0.746-1.784 | 0.41 | 0.52040 |
| **AXIN2** | rs3923087 | AA | 23(0.25) | 30(0.34) | ref |  |  |  |
|  |  | AG | 43(0.46) | 40(0.46) | 1.402 | 0.701-2.804 | 0.92 | 0.33850 |
|  |  | GG | 27(0.27) | 17(0.20) | 2.072 | 0.917-4.678 | 3.11 | 0.07794 |
|  |  | AG+GG | 70 (0.75) | 57 (0.66) | 1.602 | 0.840-3.056 | 2.06 | 0.15146 |
|  |  | A | 93(0.48) | 100(0.57) | ref |  |  |  |
|  |  | G | 101(0.52) | 74(0.43) | 1.473 | 0.972-2.233 | 3.34 | 0.06772 |
| **AXIN2** | rs3923086 | TT | 9(0.10) | 19(0.22) | ref |  |  |  |
|  |  | TG | 48(0.51) | 34(0.39) | 2.980 | 1.204-7.380 | 5.82 | 0.01581 |
|  |  | GG | 37(0.39) | 34(0.39) | 2.297 | 0.916-5.763 | 3.22 | 0.07277 |
|  |  | TG+GG | 85 (0.90) | 68 (0.78) | 2.639 | 1.122-6.204 | 5.20 | 0.02262 |
|  |  | T | 66 (0.35) | 72 (0.41) | ref |  |  |  |
|  |  | G | 122 (0.65) | 102 (0.59) | 1.305 | 0.853-1.996 | 1.51 | 0.21954 |
| **DKK3** | rs6485350 | AA | 31(0.33) | 28(0.32) | ref |  |  |  |
|  |  | AG | 48(0.51) | 40(0.46) | 1.084 | 0.560-2.100 | 0.06 | 0.81129 |
|  |  | GG | 15(0.16) | 19(0.22) | 0.713 | 0.305-1.665 | 0.61 | 0.43387 |
|  |  | AG+GG | 63 (0.67) | 59 (0.68) | 0.964 | 0.518-1.797 | 0.01 | 0.90925 |
|  |  | A | 110(0.59) | 96(0.55) | ref |  |  |  |
|  |  | G | 78(0.41) | 78(0.45) | 0.873 | 0.575-1.324 | 0.41 | 0.52165 |
| **DKK4** | rs3763511 | CC | 62(0.66) | 64(0.74) | ref |  |  |  |
|  |  | CT | 25(0.27) | 23(0.26) | 1.122 | 0.577-2.182 | 0.12 | 0.73443 |
|  |  | TT | 7(0.07) | 0(0.00) | 15.480 | 0.86-276.8 | 6.85 | 0.00885 |
|  |  | CT+CC | 32 (0.34) | 23 (0.26) | 1.436 | 0.758-2.723 | 1.24 | 0.26632 |
|  |  | C | 149(0.79) | 151(0.87) | ref |  |  |  |
|  |  | T | 39(0.21) | 23(0.13) | 1.718 | 0.979-3.017 | 3.61 | 0.05756 |
| **SFRP3** | rs288326 | AA | 1(0.01) | 2(0.02) | ref |  |  |  |
|  |  | AG | 13(0.14) | 14(0.16) | 1.857 | 0.150-22.998 | 0.24 | 0.62559 |
|  |  | GG | 79(0.85) | 70(0.82) | 2.257 | 0.2-25.43 | 0.46 | 0.49895 |
|  |  | AG+GG | 92 (0.99) | 84 (0.98) | 2.19 | 0.195-24.6 | 0.42 | 0.51501 |
|  |  | A | 15(0.08) | 18(0.10) | ref |  |  |  |
|  |  | G | 171(0.92) | 154(0.90) | 1.332 | 0.649-2.735 | 0.62 | 0.43276 |
| **SFRP3** | rs7775 | Arg | 1(0.01) | 2(0.02) | ref |  |  |  |
|  |  | Arg/Gly | 18(0.19) | 30(0.34) | 1.200 | 0.10-14.195 | 0.02 | 0.88486 |
|  |  | Gly | 75(0.80) | 55(0.64) | 2.727 | 0.241-30.841 | 0.71 | 0.39929 |
|  |  | Arg/Gly+Gly/Gly | 93 (0.99) | 85 (0.98) | 2.188 | 0.195-24.569 | 0.42 | 0.51554 |
|  |  | Arg | 20(0.11) | 34(0.20) | ref |  |  |  |
|  |  | Gly | 168(0.89) | 140(0.80) | 2.040 | 1.124-3.703 | 5.64 | 0.01753 |
| **TCF7L2** | rs12255372 | GG | 37(0.39) | 34(0.39) | ref |  |  |  |
|  |  | GT | 46(0.49) | 41(0.47) | 1.031 | 0.550-1.931 | 0.01 | 0.92410 |
|  |  | TT | 11(0.12) | 12(0.14) | 0.842 | 0.329-2.159 | 0.13 | 0.72078 |
|  |  | GT+TT | 57 (0.61) | 53 (0.61) | 0.988 | 0.544-1.796 | 0.00 | 0.96911 |
|  |  | G | 120(0.64) | 109(0.63) | ref |  |  |  |
|  |  | T | 68(0.36) | 65(0.37) | 0.950 | 0.620-1.457 | 0.05 | 0.81508 |
| **LRP6** | rs2075241 | GG | 66(0.70) | 57(0.66) | ref |  |  |  |
|  |  | GC | 28(0.30) | 27(0.31) | 0.896 | 0.474-1.693 | 0.12 | 0.73421 |
|  |  | CC | 0(0.00) | 3(0.03) | 0.124 | 0.006-2.442 | 3.38 | 0.06597 |
|  |  | GC+CC | 28(0.30) | 30(0.34) | 0.806 | 0.431-1.506 | 0.46 | 0.49881 |
|  |  | G | 160(0.85) (0.) | 141(0.81) | ref |  |  |  |
|  |  | C | 28(0.15) | 33(0.19) | 0.748 | 0.430-1.299 | 1.07 | 0.30110 |
| **LRP6** | rs2284396 | TT | 39(0.42) | 42(0.48) | ref |  |  |  |
|  |  | TC | 51(0.54) | 33(0.38) | 1.664 | 0.897-3.087 | 2.63 | 0.10510 |
|  |  | CC | 4(0.04) | 12(0.14) | 0.359 | 0.107-1.207 | 2.90 | 0.08852 |
|  |  | TC+CC | 55(0.58) | 45(0.52) | 1.316 | 0.731-2.369 | 0.84 | 0.35892 |
|  |  | T | 129(0.69) | 117(0.67) | ref |  |  |  |
|  |  | C | 59(0.31) | 57(0.33) | 0.939 | 0.604-1.460 | 0.08 | 0.77930 |

Note: OR 95% CI. Odds Ratio and 95% Confidence Interval. \* P < 0.05 was considered significant.

Similarly, rs454886 showed a strong protective effect, primarily in individuals aged 58 years or older, with both the heterozygous CT (OR = 0.330, p = 0.02) and homozygous CC (OR = 0.288, p = 0.007) genotypes associated with reduced CVD risk. Overall, both APC SNPs—rs459552 (OR = 0.494, p = 0.003) and rs454886 (OR = 0.335, p = 0.008)—uniformly showed a protective role in CVD across age groups.

Previous studies have linked rs459552 to thyroid and colorectal cancers (Figlioli et al. 2014; Rosales-Reynoso et al. 2019), while rs454886 has been associated with colorectal and breast cancers (Parine et al. 2019; Wang et al. 2008; Alanazi et al. 2013). The genetic variation in APC may modulate CVD susceptibility, possibly by regulating cardiomyocyte proliferation via asymmetric Wnt/β-catenin signaling.

### 2. AXIN2 gene

SNPs in AXIN2 showed a significant association with increased CVD risk in the study population (*Table 3*). Among the analyzed variants, rs11079571 and rs3923086 were identified as risk-associated SNPs. The association of rs11079571 with CVD was observed only in male participants (*Table 4*) and did not show evidence of age effect modification (*Tables 6 and 7*). This variant demonstrated male-specific strong risk associations, with both the homozygous minor allele (AA) and heterozygous (GA) genotypes showing elevated odds ratios (OR = 3.168 and 2.635, respectively) (*Table 4*).

In contrast, rs3923086 exhibited a significant age-dependent association, with increased CVD risk observed in individuals older than 58 years, while no gender-specific association was detected (*Tables 4–7*). For rs3923086, the heterozygous TG genotype was significantly associated with increased CVD risk in individuals aged 58 or older (OR = 2.980, p = 0.016). In addition, the TT genotype or T allele at rs3763511 was associated with a markedly increased CVD risk, particularly among male participants (OR = 9.270), suggesting a potential gender-specific genetic effect. Other AXIN2 variants (rs4791171 and rs3923087) showed no significant association with CVD. Overall, participants aged above 58 years showed a higher burden of CVD risk–associated variants, including those in AXIN2 and DKK4 (*discussed below in subtitle 3*).

Genetic variants in AXIN2 have previously been associated with a range of complex diseases, including breast and ovarian cancers and diabetic nephropathy (Crauciuc et al. 2020). Importantly, variants in AXIN genes have also been associated with congenital heart defects (CHD), with reported risk haplotypes including C–C–C and C–C–T (Crauciuc et al. 2020).

Analysis was limited to AXIN2 variants, of which two SNPs showed statistically significant associations with increased CVD risk, supporting a role for AXIN2 in CVD susceptibility. Given AXIN2’s established involvement in cardiac development and Wnt-mediated signaling, these associations suggest that age-and gender-specific genetic variation in AXIN2 may contribute to individual differences in CVD risk, potentially by modulating β-catenin signaling pathways (Loh et al. 2016).

### 3. DKK family of genes

DKK4, located on chromosome 8 (chr8:42378340), has been previously associated with colorectal and breast cancers (Al Shareef et al. 2022). SNPs in DKK family genes showed differential associations with CVD in this study. The DKK3 variant rs6485350 did not show a statistically significant association with CVD. The data showed no evidence of detectable contribution of this SNP to CVD susceptibility in the studied population.

In contrast, the DKK4-associated SNP rs3763511 demonstrated a significant association with increased CVD risk. The presence of the TT genotype or T allele was associated with higher disease risk (OR = 11.383, p = 0.004), with the effect observed predominantly in male participants (OR = 9.270, p = 0.01336) (*Tables 4 and 5*). Allelic analysis also indicated that the T allele was associated with increased CVD risk (OR = 1.588, p = 0.02013), with a dose-dependent relationship between allele frequency and disease susceptibility. Age-stratified further examination indicated that the TT genotype was enriched in individuals aged 58 years or older (*Tables 6 and 7*). Notably, participants aged 58 years or older with the homozygous TT genotype exhibited a markedly increased risk of CVD (OR = 15.480, p = 0.00885), reflecting a pronounced age-dependent association.

Genetic variation within DKK genes may therefore exert context-dependent effects, with different SNPs conferring either protective or no effects based on both gender-and age-specific association across the disease. A similar observation was previously made in lung cancer (Al Shareef et al. 2022).

### 4. SFRP3 gene

Loss or alteration of SFRP3-mediated inhibition has been implicated in multiple disease states. Variants rs288326 and rs7775 are located on chromosome 2 (positions 182,838,608 and 182,834,857, respectively) and have previously been associated with osteoarthritis and colorectal cancer, respectively (Sharma et al. 2015). Beyond oncology, altered expression of SFRP3 has been implicated in adverse ventricular remodeling and dysfunction, and experimental models suggest that elevated SFRP3 activity may improve ventricular function, reduce myocardial fibrosis, and enhance ischemic preconditioning–mediated cardioprotection by modulating Wnt/β-catenin signaling in the heart (Huang and Huang 2020).

SNPs in SFRP3 showed age-dependent associations with CVD in this study. The rs7775 variant showed a nominally significant association with increased CVD risk in individuals aged 58 years or older (OR = 2.040, p = 0.01753), with no evidence of effect modification by gender. The rs7775 Gly genotype showed the strongest association among subjects aged 58 years or older.

In contrast, rs288326, another SFRP3-associated SNP, showed a protective association in individuals older than 58 years, suggesting variant-specific, directionally distinct effects within the same gene.

Together, the observations demonstrate the presence of both risk-and protective-associated variants within SFRP3, consistent with locus-specific heterogeneity.

Collectively, the observed age-specific association of rs7775 with increased CVD risk, alongside the protective effects of rs288326, suggests that genetic variation in SFRP3 may modulate CVD susceptibility by differentially regulating Wnt signaling, particularly in older individuals.

As a part of the Wnt/β-catenin pathway, mutations in SNP rs2284396 (Genotype: TC) (OR=1.536, p=0.04523) leading to a heterozygous TC state were observed in our study, leading to an increased risk in females only (Tables *4 and 5*), and the SNP shows no correlation with age (Tables *6 and 7*). Interestingly, in our study, an LRP6 gene SNP rs2075241 (Genotype: GC; GC+CC) (OR=0.476, p=0.01546) showed a protective effect in the GC alleles in males only (*Tables 4 and 5*). This SNP showed no association with age (*Tables 6 and 7*).

### 5. LRP6 gene

In this study, two SNPs within LRP6, rs2075241 and rs2284396, located on chromosome 12 at positions 12,138,545 and 12,122,001, respectively, were evaluated. Both variants have previously been implicated in colorectal cancer, ischemic stroke, lung cancer, and cognitive impairment (Harriott et al. 2015; Parine et al. 2019; Deng et al. 2014; Alarcón et al. 2013). Notably, the rs2075241 variant has been associated with increased LDL cholesterol (LDL-C) levels, providing a plausible link between LRP6 variation and CVD risk (Alarcón et al. 2013). The LRP6 SNP rs2075241 demonstrated a protective association in the heterozygous GC genotype (OR = 0.476, p = 0.01546), as well as in the combined GC + CC genotype group (OR = 0.500, p = 0.02179), predominantly among male participants. In contrast, the LRP6 variant rs2284396 showed a nominally significant association with increased CVD risk in individuals carrying the heterozygous TC genotype (OR = 1.536, p = 0.04523), with a stronger association observed among female participants (OR = 2.082, p = 0.02279) (*Table 5*).

In the present cohort, rs2284396 showed a nominally significant association with increased CVD risk in females only, with individuals carrying the heterozygous TC genotype exhibiting elevated odds of disease (OR = 1.536, p = 0.04523). In contrast, rs2075241 demonstrated a protective association, with the heterozygous GC genotype associated with reduced CVD risk (OR = 0.476, p = 0.01546). The results indicate variant-specific and directionally distinct associations within the LRP6 locus.

Consistent with prior reports, LRP6 has been linked to adverse cardiac outcomes, including impaired cardiac homeostasis and arrhythmogenesis. Reduced LRP6 expression has been associated with arrhythmias, and a study in a Han Chinese population reported that LRP6 variants increased the risk of sudden cardiac death in patients with chronic heart failure (Guo et al. 2021). Taken together, our results support a role for LRP6 genetic variation in affecting cardiovascular disease susceptibility, likely through combined effects on lipid metabolism and canonical Wnt/β-catenin signaling.

### 6. TCF7L2 gene

Genetic variation in TCF7L2 has been extensively associated with type 2 diabetes mellitus, a major metabolic risk factor for CVD. The rs12255372 polymorphism, located on chromosome 10 at position 113,049,143, has previously been reported to be associated with diabetes mellitus and CHD (Muendlein et al. 2011). Moreover, interactions between this variant and high-density lipoprotein cholesterol (HDL-C) have been suggested to influence cardiovascular disease risk (Bodhini et al. 2017). Consistent with the results, several common variants in TCF7L2 have been implicated in type 2 diabetes susceptibility, indirectly linking this locus to cardiovascular disease risk through the metabolic pathways (Del Bosque-Plata et al. 2021).

However, in the present study, no statistically significant association was observed between TCF7L2 rs12255372 and CVD. This suggests that, despite its established role in metabolic regulation and Wnt signaling, TCF7L2 variation may not independently contribute to CVD susceptibility in this cohort.

### 7. Other genes

No statistically significant associations were observed between CVD and CTNNB1 (β-catenin) SNPs rs13072632 and rs4135385, AXIN2 SNPs rs4791171 and rs3923087, DKK3 SNP rs6485350, or TCF7L2 SNP rs12255372, indicating a lack of detectable association for these variants in the present study cohort. Consistent with this, the DKK3 variant rs6485350 did not demonstrate susceptibility to CVD.

## Discussion

This study reveals that genetic variation within the canonical Wnt/β-catenin signaling pathway influences CVD risk in a highly context-dependent manner, with effects shaped by both age and sex. Protective associations were observed for variants in APC (rs459552 and rs454886) and LRP6 (rs2075241), particularly among male participants and within specific age groups, suggesting a potential role for these variants in lowering CVD susceptibility. In contrast, increased disease risk was linked to variants in AXIN2 (rs11079571 and rs3923086), DKK4 (rs3763511), SFRP3 (rs7775), and LRP6 (rs2284396), with their effects varying according to age and sex. Together, these findings highlight the complex and finely balanced role of Wnt signaling in cardiovascular pathology.

Not all variants within this pathway contributed to disease risk. Several loci, including variants in CTNNB1, TCF7L2, DKK3, and selected AXIN2 SNPs, showed no detectable association with CVD, underscoring the highly variant-specific nature of genetic risk within the Wnt pathway. Importantly, both intracellular regulators of Wnt signaling (APC, AXIN2, and LRP6) and extracellular modulators (DKK4 and SFRP3) emerged as contributors to disease susceptibility, emphasizing the multi-level regulation of this pathway in cardiovascular health.

While these results provide valuable insight, they should be interpreted in light of potential limitations, including unmeasured environmental and lifestyle factors that may influence cardiovascular risk. Future studies incorporating detailed lifestyle, metabolic, and familial data, along with larger and ethnically diverse cohorts, will be essential to validate these findings and to explore their potential application in cardiovascular risk prediction and precision medicine.

In conclusion, this study shows that genetic variation across multiple regulatory levels of the canonical Wnt/β-catenin signaling pathway contributes to cardiovascular disease (CVD) susceptibility in a context-dependent manner. Both protective and risk-associated variants were identified in intracellular regulators (APC, AXIN2), the Wnt co-receptor (LRP6), and extracellular modulators (DKK4, SFRP3), highlighting the importance of coordinated Wnt signaling control in cardiovascular health. Variants in APC and AXIN2 may influence β-catenin turnover and transcriptional activity, thereby affecting cardiomyocyte proliferation, hypertrophic responses, and myocardial remodeling. Polymorphisms in LRP6 suggest a mechanistic link between Wnt signaling, lipid and glucose metabolism, and cardiovascular risk, with notable age-and sex-specific effects. Similarly, variation in extracellular Wnt modulators such as DKK4 and SFRP3 is likely to alter ligand availability and signaling amplitude, processes that influence fibroblast activation, extracellular matrix deposition, and myocardial fibrosis, particularly in older individuals.

Although not all Wnt pathway components showed significant associations, the overall pattern supports a polygenic, network-based model in which subtle dysregulation at multiple signaling nodes collectively shapes cardiovascular risk. Importantly, CVD susceptibility extends beyond single-nucleotide polymorphisms and involves a broad spectrum of genetic variation, including copy number changes, insertions and deletions, rare coding mutations, regulatory variants, mitochondrial alterations, and epigenetic modifications. These genetic factors converge on interconnected pathways—most notably Wnt/β-catenin, TGF-β-mediated fibrotic signaling, and metabolic regulation—to influence cardiomyocyte survival, fibrotic remodeling, vascular homeostasis, and energy metabolism. Future studies integrating functional genomics with longitudinal clinical and lifestyle data, and conducted in larger and more diverse populations, will be essential to validate these findings and to define their translational potential for CVD risk stratification and therapeutic targeting.

Protein-level analysis adds important biological insight into the genetic associations observed in this study. Missense variants such as APC rs459552 (Val1822Asp) and SFRP3 rs7775 (Arg324Gly) directly alter amino-acid sequence and are therefore the most likely to have functional effects. The Val1822Asp substitution in APC occurs within the AXIN-binding region that regulates β-catenin degradation and is predicted to subtly adjust, rather than disrupt, Wnt/β-catenin signaling. This fine-tuning may help limit maladaptive profibrotic signaling while preserving essential cardiac repair processes, providing a plausible explanation for its protective association with cardiovascular disease. Similarly, the Arg324Gly change in SFRP3 is likely to weaken extracellular inhibition of Wnt ligands, promoting age-dependent activation of profibrotic pathways. In contrast, most variants identified in AXIN2, LRP6, CTNNB1, and TCF7L2 are non-coding and likely influence cardiovascular risk through regulatory effects on gene expression rather than direct protein disruption. Together, these findings highlight a polygenic model in which both coding and regulatory variation across the Wnt/β-catenin pathway converge to shape ventricular remodeling and cardiovascular disease susceptibility.

From a clinical perspective, these findings highlight the complex and polygenic nature of cardiovascular disease. Rather than being driven by single genetic changes, CVD risk appears to arise from the combined effects of multiple variants acting across interconnected pathways. This supports the value of integrating multi-variant genetic profiling into cardiovascular risk assessment, particularly for individuals with early-onset, familial, or otherwise unexplained disease. Advances in genomic technologies offer an opportunity to identify functionally relevant variants within key signaling pathways, enabling more precise risk stratification. As our understanding of these genetic contributions improves, it may also inform the development of targeted and pathway-guided therapeutic strategies, ultimately supporting more personalized approaches to cardiovascular disease management.

## Data Availability

All data produced in the present work are contained in the manuscript

